# Identifying regions for enhanced control of *gambiense* sleeping sickness in the Democratic Republic of Congo

**DOI:** 10.1101/2020.07.03.20145847

**Authors:** Ching-I Huang, Ronald E Crump, Paul E Brown, Simon E F Spencer, Erick Mwamba Miaka, Chansy Shampa, Matt J Keeling, Kat S Rock

**Author notes:** these authors contributed equally to this work (Ching-I Huang, Ronald E Crump).

## Abstract

*Gambiense* human African trypanosomiasis (sleeping sickness, gHAT) is a disease targeted for elimination of transmission (EOT) by 2030. Despite the number of new cases reported annually being at a historical minimum, the likelihood of achieving EOT is unknown. We utilised modelling to study the impact of four strategies comprised of currently-available intervention methods including active and passive screening and vector control (VC) on transmission across 168 health zones in the Democratic Republic of the Congo. By estimating the median year of EOT and the probability of EOT by 2030 under each strategy, the model predicts only 81 health zones are on track to achieve the EOT target using medical-only strategies and this number drops to 52 when uncertainty is considered (*>* 90% probability). Although all health zones are predicted to meet EOT by 2030 under strategies with VC, blanket coverage is impractical so this analysis provides a priority list of health zones for consideration for supplementary VC implementation in conjunction with medical interventions.

## Introduction

The pinnacle of success for an infectious disease programme is to drive the disease to eradication, resulting in complete removal of morbidity and mortality, yet no longer requiring interventions. Of the human diseases targeted for eradication, only one – smallpox – has currently achieved this objective, yet there are several for which this remains the (potentially illusive) goal (such as polio, Guinea worm, and yaws). Clear lessons that can be learnt from many eradication programmes are: (i) the often slow progress from low to very low case burden, (ii) the ever-increasing effort required per case to tackle remaining infection, and (iii) the question of whether eradication is even epidemiologically or operationally feasible.

One step down from eradication is elimination of transmission (EOT) to humans, acknowledging that transmission pockets could persist in non-human animal cycles. This goal, arguably, may be almost as challenging and fraught with the same hurdles to overcome as eradication itself. *Gambiense* human African trypanosomiasis (gHAT, sleeping sickness) is one such disease with the EOT goal and within the last decade it was still known to be extant in 15 countries in West and Central Africa^1^. This parasitic infection is transmitted to humans via bites by tsetse, with gHAT symptoms typically increasing in severity over several years and leading to death without treatment. The remarkable progress made to bring down the case burden across the continent – falling to below 10,000 cases in 2009 for the first time since the most recent epidemic started in the 1970s, and to only 953 in 2018^1–3^ – has sparked optimism that EOT may be possible and the World Health Organization (WHO) has set the goal of EOT by 2030^4,5^. Indeed gHAT fulfils some of the criteria associated with an “eliminable” disease^6^: we have a range of field-proven tools and associated delivery mechanisms as well as means of diagnosis and surveillance. Unlike smallpox, gHAT is not vaccine-preventable, but wide-spread testing, diagnosis, and treatment have worked well to curtail transmission. The key question is whether current tools for gHAT are sufficient to reach EOT in the next ten years, and if so, how expansive might their use have to be to get there.

The Democratic Republic of the Congo (DRC) is the country with the most reported gHAT cases. Due to the concerted efforts of the national sleeping sickness control programme in DRC (PNLTHA-DRC), the number of reported cases dropped below 1,000 in the country in 2018. However, the DRC still accounted for nearly 70% of global cases (660 out of 953 cases) in that year^1,3^. Therefore, the DRC is the most critical country on which the achievement of EOT by 2030 hinges.

In order to assess EOT feasibility, this study focuses on quantitative forecasting of gHAT across the endemic health zones (each have a population of around 100,000 people) in DRC to examine if, how, and when EOT could be expected under strategies based on currently available tools. Previous DRC-specific predictive modelling studies have provided insights into expected timelines to EOT in Equateur province^7^, and parts of Bandundu province^8–12^ under continuation of medical-based strategies with or without vector control. From these studies it is clear that a one-size-fits-all approach is unlikely to be sufficient to meet this highly ambitious target in the next decade. Although coverage of active screening has been driven by local numbers of cases, additional data-driven guidance could help to further tailor strategy selection.

In this article, we enlarge the geographical scope of previous predictions to include 168 endemic health zones across the whole country, utilising the results of previous fitting^13^ to examine the strategies of active screening (AS) with or without supplemental, large-scale vector control (VC) on top of the local passive screening (PS) system to stop gHAT transmission by 2030 in DRC. A graphical user interface (GUI) to complement this article was set up to provide full model outputs. In this analysis we aim to identify regions which are likely to be successful in achieving local EOT on their current trajectory, and ones where enhanced control may be required to meet this target. Furthermore, we provide a priority list of health zones where intensification of strategies is most urgent based on past intervention coverage and projected timelines to EOT.

## Results

### Projection trends in different risk settings

We used our previously fitted, deterministic SEIRS-type model of gHAT to simulate forward projections in 168 health zones under four strategies: two medical-only strategies which comprise of active and passive screening (MeanAS and MaxAS), and two medical strategies with supplemental vector control (MeanAS+VC and MaxAS+VC). The active screening coverage is either the mean of the last five years of data (2012–2016) or the maximum level achieved historically (2000–2016). Projections were run for 2017–2050 and incorporated parameter uncertainty. Table 1 in the Methods gives more detailed information on the strategies. Non-endemic health zones, and those with little intervention, and/or case reporting were excluded from the original model fitting and hence from these projections^13^.

**Table 1.**
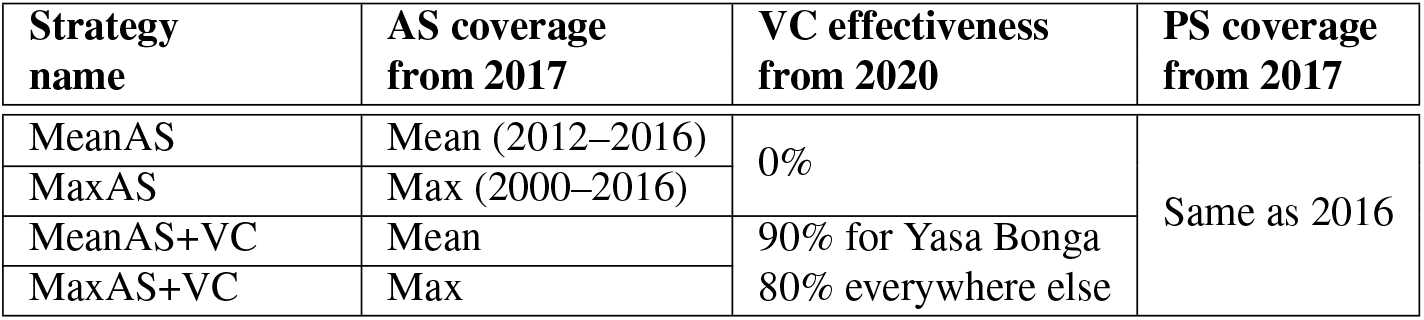
Strategies considered for projections (2017–2050). VC effectiveness is determined by the proportional reduction in tsetse population after one year of implementation. Strategies without VC are not considered in Yasa Bonga because VC has been implemented since the middle of 2015.

Figure 1 shows assumed numbers of people screened and model outputs (i.e. active and passive cases, new infections, and probability of EOT) for the four strategies in two example health zones: Kwamouth (former Bandundu province) and Tandala (former Equateur province). Both health zones had significant numbers of cases in the early 2000s and still have on-going transmission despite annual AS. Kwamouth, with 1068 reported cases in 2012–2016 (estimated 2015 population of 127,205), falls within WHO’s definition of a “high-risk” category for gHAT (1–10 cases/1,000 per year averaged over five years), while Tandala is only “low-risk” (38 reported cases in 2012–2016 and estimated 2015 population of 274,945 – i.e. 1–10 cases/100,000 per year). Historical AS data shows that Kwamouth had substantially higher proportions of people screened than Tandala. Despite very high coverage of AS in Kwamouth, achieving EOT by 2030 is predicted to only be possible when VC is added – the model suggests that transmission will be interrupted completely within four years once VC begins. Unlike Kwamouth, Tandala appears extremely likely to achieve EOT by 2030 with the MaxAS strategy and EOT even occurs in 60% of projections under the less intensive MeanAS strategy. Projections under each strategy for each health zone can be found in our graphical user interface (URL given in Graphical user interface (GUI) section, below).

**Figure 1.**
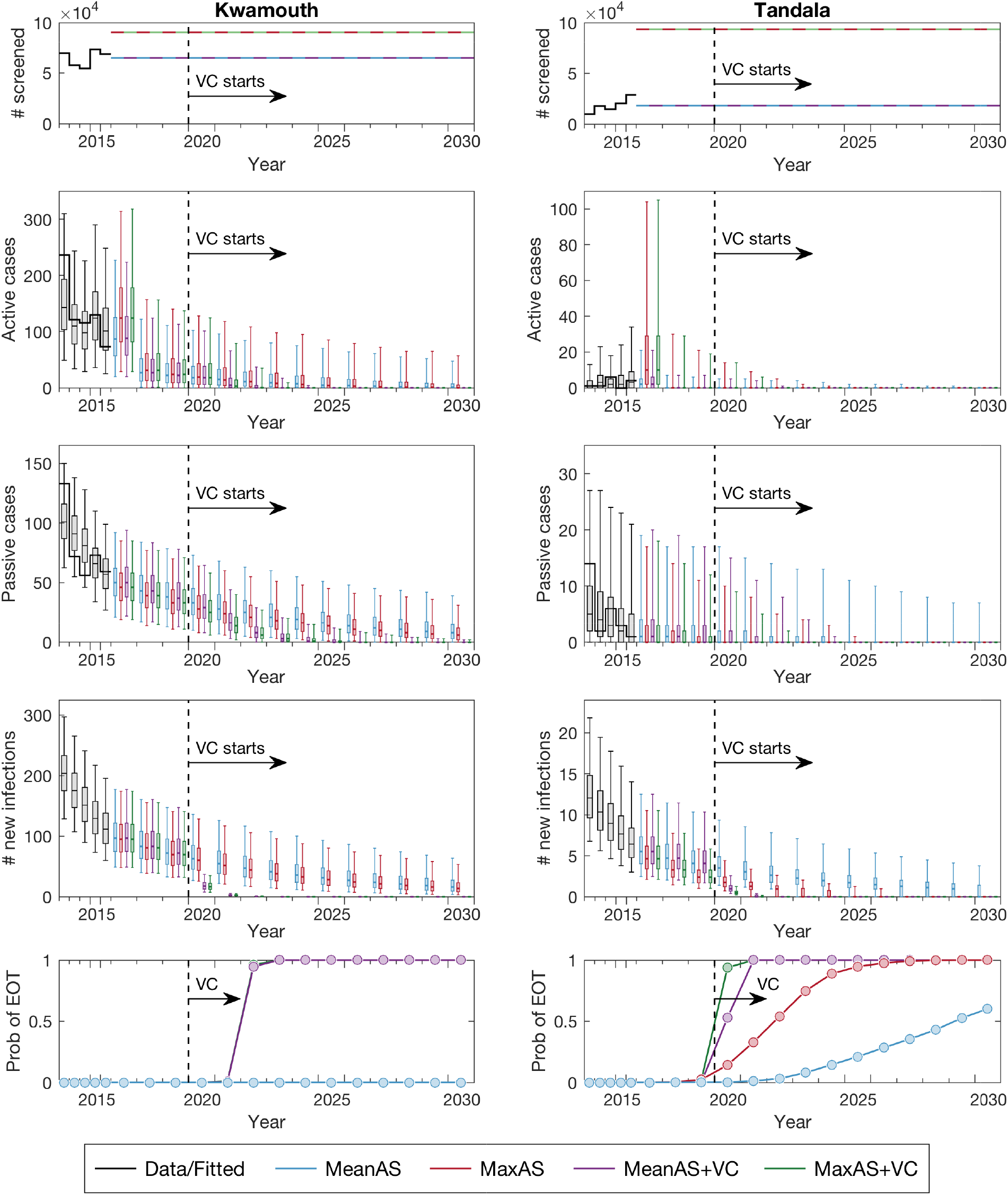
Time series of key model outputs in two example health zones. Kwamouth (left panels) in former Bandundu province and Tandala (right panels) in former Equateur province represent a high-risk and a low-risk health zone, respectively. The top row shows the number of people actively screened, the middle rows show three direct model outputs (active cases, passive cases and underlying new infections from top to bottom), and the bottom row shows the probability of achieving EOT by year. Black lines and box plots indicate data and model fits in the last five years (2012–2016), coloured dashed lines denote the assumed AS starting in 2017, and colour box plots and circles present the predictions for four strategies (as defined in Table 1). Box plots with whiskers showing 95% prediction intervals summarise parameter and observational uncertainty. Full model outputs (2000–2050) of all 168 analysed health zones are available in the GUI).

### Timelines to, and certainty of, EOT

The year of elimination of transmission (YEOT) is defined as the first year that the EOT criterion (i.e. number of new infections is less than one) is met. Health zone maps of the median YEOT under the four strategies are shown in Figure 2. It is possible for different strategies to have very similar YEOT distributions within a health zone if EOT is expected to have already occurred. Using the median value of YEOT, health zones can be classified into three categories: on track (YEOT ≤ 2030), slightly behind schedule (2030 < YEOT ≤ 2040), and greatly behind schedule (YEOT > 2040) to meet the EOT goal. We predict 74 health zones are on track, 29 are slightly behind schedule, and 65 are greatly behind schedule under the MeanAS strategy. An extra seven health zones are on track while 62 remain greatly behind schedule under the MaxAS strategy. Data shows low coverage of AS (defined as lower than 25% for mean coverage and 40% for maximum coverage) may be responsible for predicted delays in EOT in health zones outside the former Bandundu province. With VC starting in 2020, all health zones are predicted to achieve EOT by 2024. The MaxAS+VC strategy could further bring forward YEOT by up to one year (although the five year data bins in Figure 2 obscure this nuance).

**Figure 2.**
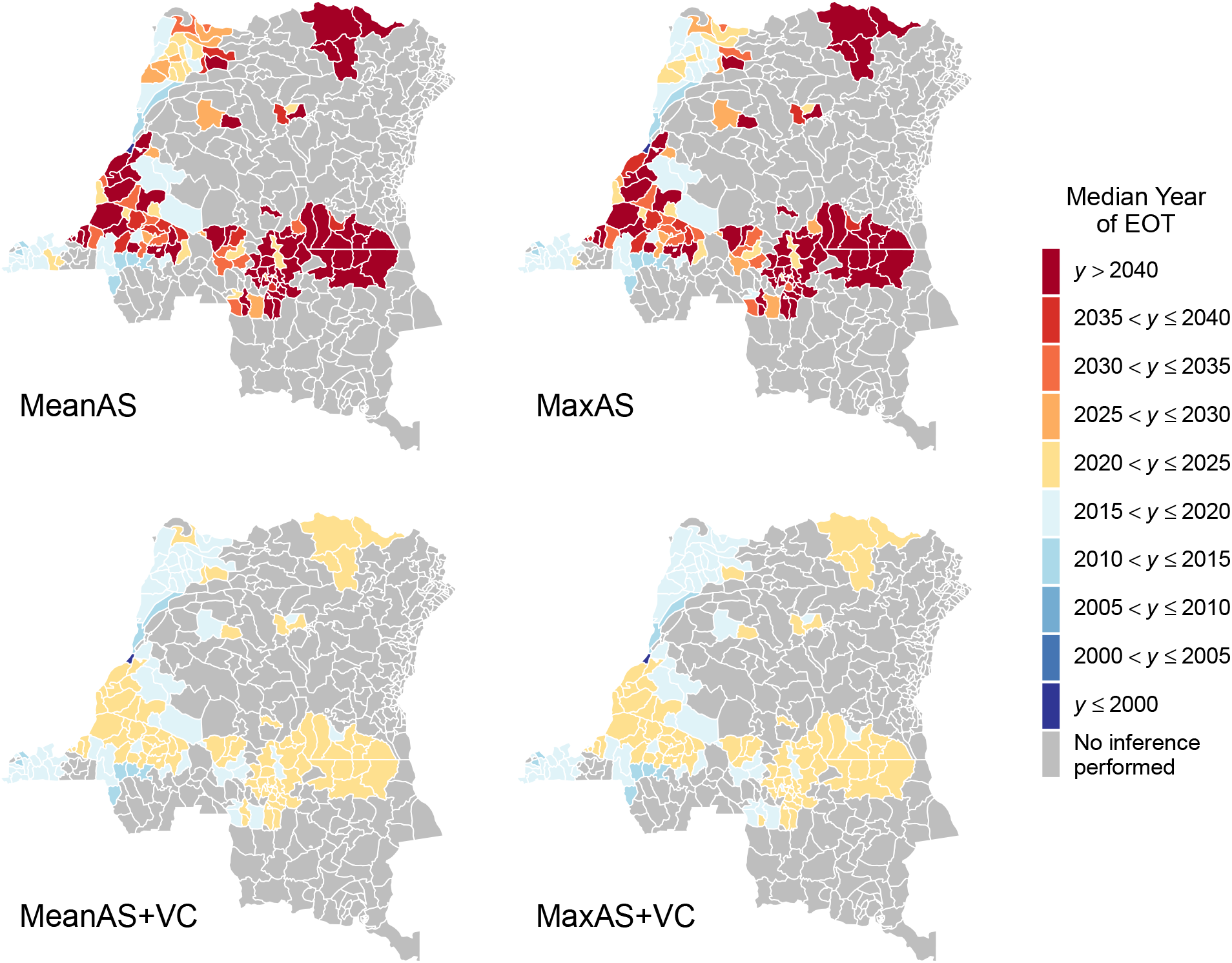
Health zone median year of elimination of transmission (YEOT) maps for the DRC. The median YEOT provides the year in which 50% of model simulations reach EOT in each health zone. The top two maps show strategies without VC (except for Yasa Bonga health zone which is shown with VC in all maps) and the bottom maps have VC strategies with 80% vector reduction. The left maps simulate continuation of the mean AS coverage and the right two simulate maximum AS coverage. The uncertainty of YEOT is not shown in these maps (only the average prediction). The exact median values and 95% prediction intervals for YEOT are available in the GUI).

The median YEOT provides a point estimate of when to expect EOT but not the degree of certainty that the goal will be met by 2030. The probability of elimination of transmission (PEOT) by 2030, which reflects the distribution of YEOT, captures the uncertainty of model predictions. Consequently, low values of median YEOT cannot guarantee EOT by 2030. One example is Inongo in the former Bandundu province, which has a median YEOT of 2019 but the PEOT by 2030 is < 1 under both the MeanAS and MaxAS strategies. Figure 3 shows PEOT by 2030 in each health zone under four strategies. Three uncertainty categories of model predictions are particularly interesting: EOT is very likely to be met by 2030 if PEOT > 0.9, EOT by 2030 is highly uncertain when 0.3 < PEOT < 0.7, and EOT is very unlikely to be met if PEOT < 0.1. The model predicts 42 health zones are very likely to meet the goal and 61 are almost certain to miss it under the MeanAS strategy. High uncertainty in EOT is reported in 33 health zones. Despite the distribution of YEOT being shifted forward under the MaxAS strategy, only ten extra health zones become very likely to meet the goal by 2030 while 24 health zones remain highly uncertain because of their wide YEOT distributions. With VC starting in 2020, a tight distribution of YEOT means EOT by 2030 is extremely likely everywhere even if its median is quite close to 2030.

**Figure 3.**
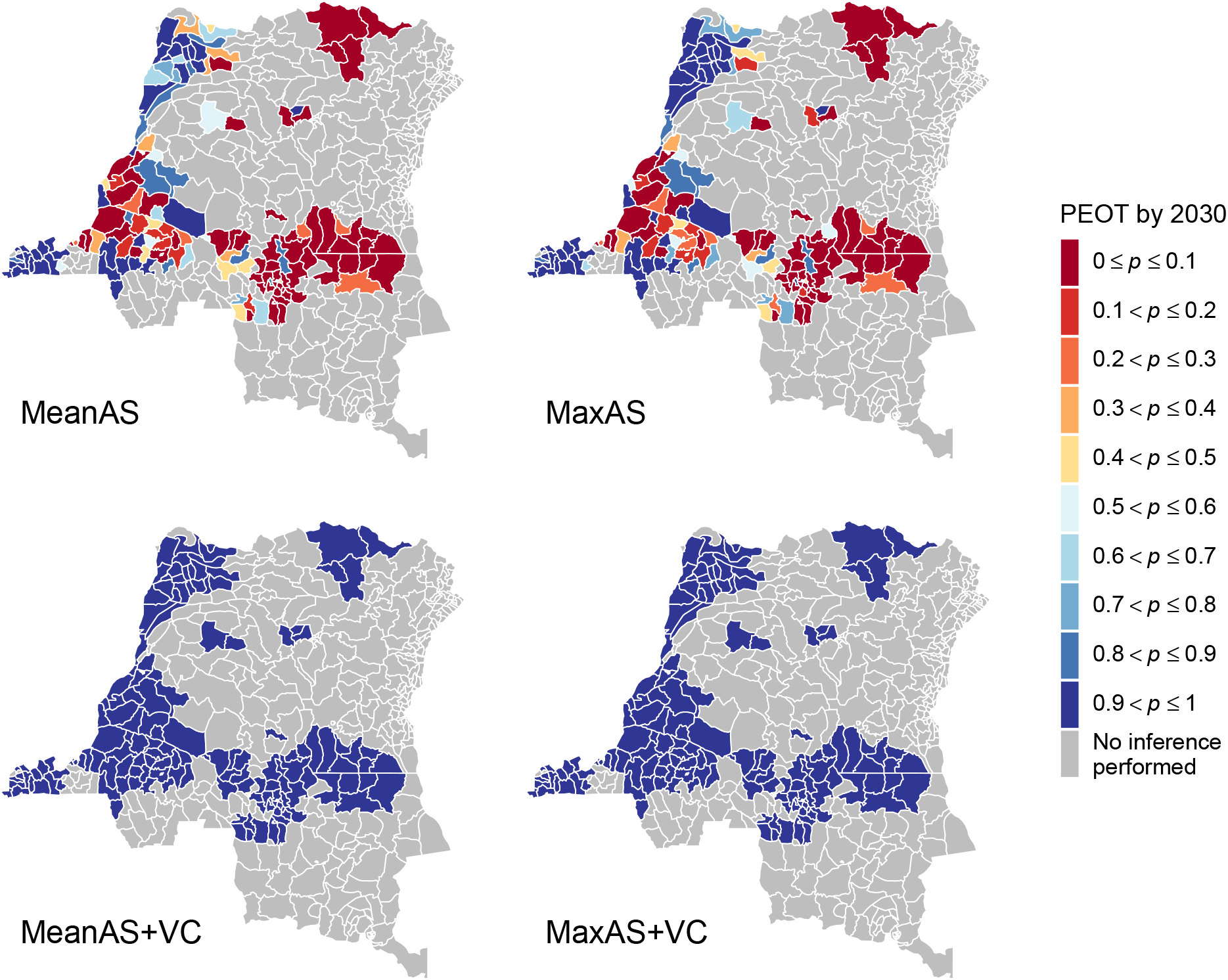
Health zone probability of elimination of transmission (PEOT) by 2030 maps for the DRC. PEOT reveals the uncertainty of model predictions about whether EOT will occur. Health zones with PEOT > 0.9 (dark blue) will be very likely to achieve EOT by 2030, and PEOT < 0.1 (dark red) will be very unlikely to meet it. Health zones with mid-range PEOT (0.3–0.7) have high uncertainty in the success or failure of the strategy to meet the goal either because (1) the median YEOT is close to 2030, or (2) the wide distribution in the predicted YEOT. The two identical maps (with PEOT = 1 everywhere) at the bottom show that VC is an efficient tool which ensures EOT has extremely high certainty. Maps of PEOT by other years are available in the GUI).

### Prioritising health zones

Decision-making for gHAT strategy is challenging; national programmes have the flexibility to implement nuanced, spatially-heterogeneous interventions, however they must adhere to more general WHO recommendations and budget constraints. In the present study we rank strategies by how ambitious the use of additional interventions is and examine the minimum required to meet the 2030 EOT goal in each health zone – referred to here as the “preferred strategy”. Maps showing the preferred strategy under different levels of certainty in EOT as predicted by the model are given in Figure 4. Under the criterion of PEOT > 0.9 (left map), preferred strategies are defined as the strategies which achieve EOT by 2030 in at least 90% of simulations. The criterion of PEOT = 1 (right map) further restricts preferred strategies to achieve the goal by 2030 in all simulations. According to the ordered ranking (MeanAS, MaxAS, MeanAS+VC, and MaxAS+VC), the least ambitious strategy among all that meet the PEOT criterion is selected as the preferred strategy. This order of ambition ranking was based on the following principles: MeanAS represents the continuation of current intervention, MaxAS is the highest level of intervention implemented to date, and VC is a new intervention to all health zones except Yasa Bonga. Notably MaxAS+VC is absent in any of the preferred strategy maps because all health zones are expected to achieve the EOT goal by 2030 under MeanAS+VC strategy which requires less resources. Maps showing lower PEOT thresholds (from PEOT = 0.5 upwards) can be found in the GUI (URL given in Graphical user interface (GUI) section, below) and far less intensification would be required if a 50% probability of meeting the goal by 2030 is considered to be sufficient.

**Figure 4.**
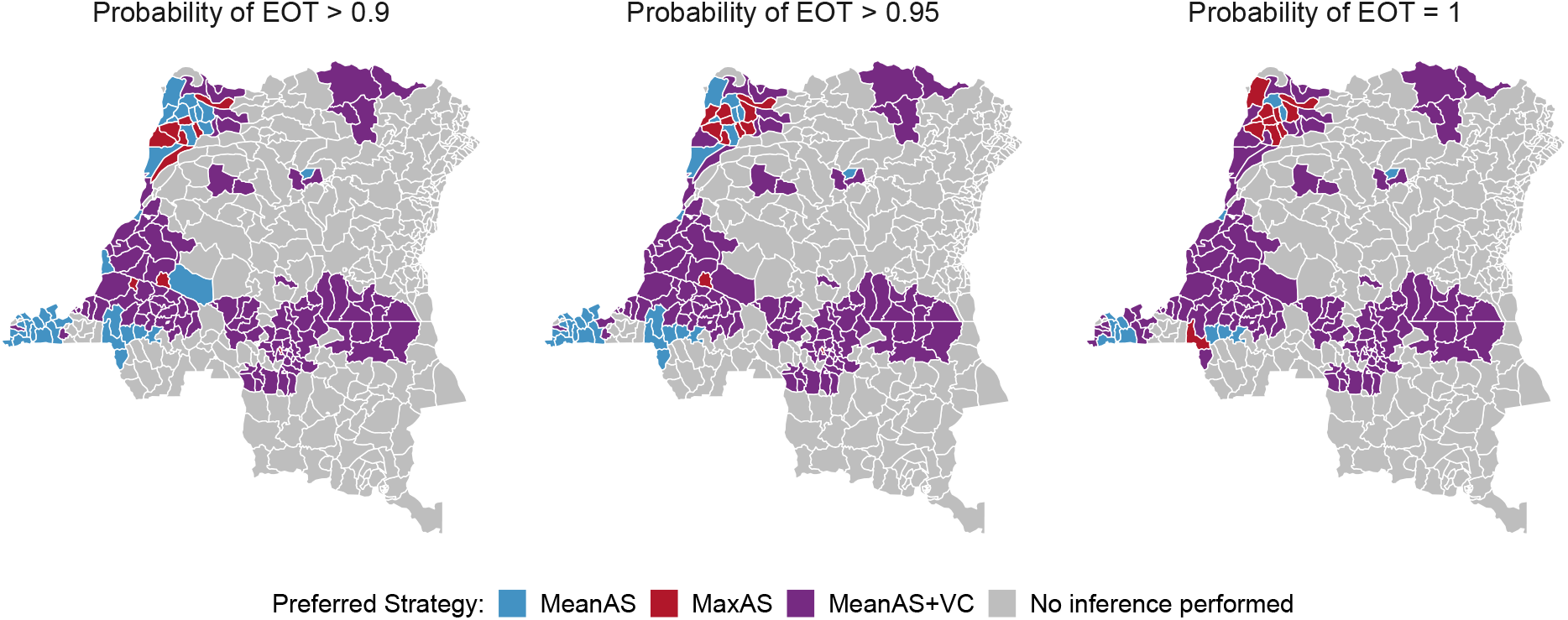
Health zone preferred strategy maps for EOT by 2030 in the DRC. The preferred strategy is defined as the least ambitious strategy which is predicted to achieve EOT by 2030 with a prescribed confidence level (90%, 95% and 100%). The order of ambition ranking is MeanAS, MaxAS, MeanAS+VC and MaxAS+VC. All health zones are predicted to achieve EOT by 2030 (PEOT = 1) under MeanAS+VC strategy so MaxAS+VC is absent here. MeanAS and MaxAS strategies were not considered in Yasa Bonga because VC started in mid-2015. Preferred strategy maps for smaller PEOT thresholds are available in the GUI.

Switching to intensified strategies is generally expected to increase confidence that EOT will be achieved, although the model predicts a relatively low percentage of health zones will eliminate transmission by medical-only strategies with high probability (31% under PEOT > 0.9 and 17% under PEOT = 1). We used the historical data in conjunction with model assumptions to understand the causes of the apparent high need for VC and suggest where and what kind of intensified interventions could result in the achievement of EOT by 2030.

Using the WHO’s risk categories, health zone can be classified as: moderate- or high-risk (≥ 1 case per year on average per 10,000), or low- or very low-risk (≥ 1 but < 100 cases per year on average per 1,000,000). Based on data from 2012–2016, there are 125 health zones in low- or very low-risk categories. On may assume these health zones should be on track to meet EOT by 2030 since they have low reported cases in recent years, however, the model predicts the majority of them (87 health zones) need VC to achieve EOT by 2030 with more than 95% probability. The apparent discrepancy comes from large uncertainty in model predictions due to limited information arising from low AS coverage. AS provides precious information on quantifying the underlying transmission and affects model predictions. In order to maximise resource efficiency, reductions in AS commonly happen when fewer cases are reported. More than 95% of the low- or very low-risk health zones screened a total of less than 50% of its population in the last five years (i.e. less than 10% annually). As a result, VC is favored in model predictions due to lack of information and may be unnecessary in practice in low- or very low-risk health zones. For moderate- or high-risk health zones, the model predicts nearly all health zones (37 out of 43) need VC to meet EOT by 2030 with more than 95% probability. Although VC seems a reasonable tool in moderate- or high-risk health zones, unfortunately it is unlikely to be practical to roll out large-scale VC in all of them in this short timeframe. In health zones outside former Bandundu province that are greatly behind schedule the maximum observed coverage was 40%. By identifying health zones which have had high AS coverage (using this threshold) but also have low probability of meeting EOT by 2030 we compiled a priority shortlist of health zones where VC is highly recommended by mathematical modelling: Bagata, Bandundu, Bolobo, Kikongo, Kwamouth, and Masi Manimba all in the former Bandundu province (as shown in Figure 5). More than 95% of health zones have mean AS coverage lower than 25%, therefore a secondary suggestion is to increase the coverage of AS to at least 25% especially in the moderate- or high-risk health zones.

**Figure 5.**
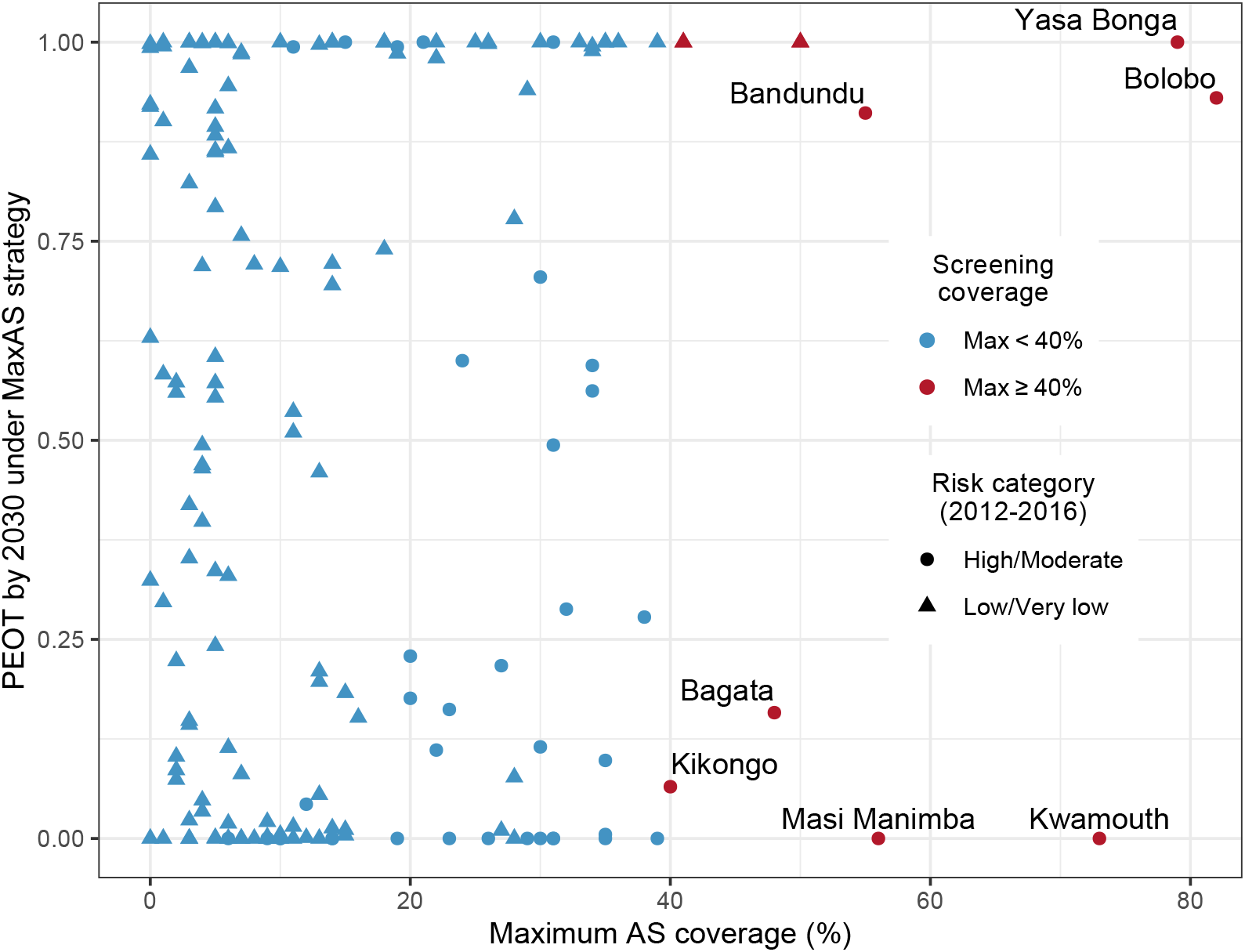
Identified health zones for supplementary VC implementation. The risk category of each health zone is defined based on data from 2012–2016 according to the thresholds defined by the WHO. High- or moderate-risk health zones are represented by coloured circles. Low- or very low-risk health zones are represented by coloured triangles. Colours indicate the coverage of maximum AS; blue denotes health zones with low maximum AS coverage (< 40%) and red denotes health zones with high maximum AS coverage (≥40%). A cutoff of 40% on maximum AS coverage based on data analysis in YEOT and PEOT is introduced to differentiate health zones of good screening coverage from moderate-to-poor screening coverage. PEOT by 2030 under MaxAS strategy represents the maximum probability of EOT with the highest level of intervention implemented to date. Our model identifies a priority list of six health zones (highlighted by their names, Bagata, Bandundu, Bolobo, Kikongo, Kwamouth and Masi Manimba in the former Bandundu province) for consideration for supplementary VC implementation because these health zones have less than 95% chance of achieving EOT under good AS coverage. N.B. Yasa Bonga is highlighted for VC implementation because implementation started in mid-2015.

### Graphical user interface (GUI)

A graphical user interface (GUI), hosted at https://hatmepp.warwick.ac.uk/projections/v1, was built to provide interactive visualisation of the data and model outputs of all 168 analysed health zones. The time series figures including the number of people actively screened, active cases, passive cases, and underlying new infections are re-generated automatically when a health zone is selected from the drop-down menu. The predicted elimination map shows a graphical summary of YEOT and PEOT. The exact median values and 95% prediction intervals for YEOT are available when hovering over health zones on the map. While the map defaults to PEOT by 2030, maps of PEOT by other years (2020–2040) are available via a controlling slider. In a separate tab, the preferred strategy map is shown and defaults to a threshold of PEOT > 0.9. Preferred strategy maps for lower PEOT thresholds are available by selecting the desired PEOT level via the controlling slider.

## Discussion

The presented model predictions are based on fitting the model to 2000–2016 case data^13^ and assumptions of different strategies starting from 2017. This is the first publicly available analysis of gHAT predictions for health zones across the whole of DRC, and highlights regions we expect to be successful, and those where there may be challenges in achieving the WHO 2030 target of EOT. By providing both average predictions, 95% prediction intervals for when we expect EOT to be met (available in the GUI), and also the probability of meeting the goal by 2030 in each health zone, we aim to quantify not only regions which may need intensified strategies, but those where current data may not be sufficient to generate predictions with high certainty.

This model utilises a set of parameters that achieve good health zone-level fits to the WHO HAT Atlas data^3,14,15^ aggregated by health zone and year. Without taking into account the potential impacts of animal reservoirs, asymptomatic infections, and secondary infections from host movement, our model results could be optimistic on the issue of EOT. However, our predicted regions for enhanced control will remain on the list should a these alternative model formulations be utilised in future modelling exercises. Given that our predictions are possibly optimistic, yet we still find health zones unlikely to meet the 2030 EOT goal, this highlights the urgent need to strengthen interventions in these locations.

The preferred strategy maps in Figure 4 show that the MeanAS+VC strategy is needed in a large proportion of the health zones. This finding brings up a serious concern about the feasibility of scaling up VC in order to achieve EOT in ten years under resource constraints. The implementation of VC began in the southern part of Yasa Bonga in 2015 and expanded to cover three large rivers (Lukula, Kafi, Inzia) and many of the tributaries linked to fish ponds by 2017^16^. Scaling up of VC was slow and its feasibility was mainly determined by the availability of financial and human resources for this new intervention. The integration of data, model assumptions, and model predictions identifies a priority shortlist of six health zones; Bagata, Bandundu, Bolobo, Kikongo, Kwamouth, and Masi Manimba in the former Bandundu province (all of which have maximum AS coverage ≥ 40%) as regions where VC is predicted to be a necessary supplementary tool for eliminating transmission by 2030. Comparing our priority list for VC to the planned VC scale-up in DRC guided by recent case data, modelling and habitat suitability (https://www.lstmed.ac.uk/projects/tryp-elim-bandundu), 5 of 6 priority health zones identified by our model are targeted as operational areas for VC rollout. Furthermore, our model suggests that the other health zones earmarked for scale-up (including Bulungu, Bokoro and Yumbi) would have been unlikely to meet EOT by 2030 (<60%) without VC interventions.

Other health zones predicted to miss the 2030 EOT goal could also benefit from this tool, although careful consideration is required to assess whether scaling up medical interventions is easier to implement than introducing large-scale VC. The reported effectiveness of VC is high in general but the variations between locations are non-negligible. According to our sensitivity analysis on the effectiveness of VC (Supplementary Fig. 1), the time difference in achieving EOT could be several years longer with only 60% annual tsetse reduction, but this is still substantially faster than with medical-only interventions in many settings. Our model forecasting would be more accurate if the location-specific effectiveness of VC – which remains unknown in most health zones – was taken into account. Pessimistic model predictions can be found in some health zones where the coverage of AS is very low recently or historically. Low AS coverage creates additional uncertainty in model outputs and therefore can make model predictions overly pessimistic (i.e. they could overstate the need for VC in low- or very low-risk health zones). Exploring the minimum AS coverage required to achieve EOT by 2030 would be another mathematical modelling approach to address where and what kind of intensified interventions are needed to achieve EOT.

When the new data from 2017 onward becomes available, we will be able to use it to validate our model by changing assumed AS coverage to actual numbers of people screened and then comparing the predicted active and passive cases to reported cases. Subsequent re-fitting to the recent case data would further refine model predictions presented here and is an important step in the continuous process of modelling to support policy under NTD-PRIME principles^17^ (Supplementary Table 3). Our model framework is flexible and could be used to predict the impact of unexpected future changes by estimating how they could alter observable (i.e. reported cases and deaths) and unobservable variables (i.e. new infections); in the present climate of the COVID-19 pandemic and recent Ebola outbreaks in gHAT-endemic parts of DRC, this is particularly relevant and could provide support in planning whether subsequent gHAT interventions should be altered due to unforeseen interruptions.

The impact of other factors such as the screening of high-risk populations and the presence of animal reservoirs on gHAT transmission have been studied by mathematical modelling^8,9,18–21^. Recruiting high-risk individuals can, unsurprisingly, improve the effectiveness of AS and bring down the YEOT substantially^9^; the present framework could be extended to quantify the impact of this type of improved AS. Models considering an animal reservoir have largely been inconclusive about the presence of zoonotic transmission (when fitted to longitudinal human case data) however they have indicated that animal reservoirs are unlikely to maintain the infection by themselves^8,19^. An analysis including animal reservoirs could yield different results for YEOT predictions presented here, although our previous work suggests that we would probably not expect large qualitative differences. Another concern is that transmission could be maintained through asymptomatic humans^22,23^. Although a few modelling studies have utilised frameworks explicitly incorporating asymptomatic human infections^20,21,23^, there is limited observational data to parameterise them with high certainty, and it is unclear how their inclusion in this study would impact projections. Host movement may play some role in spreading diseases, which not only makes EOT harder to achieve but also could potentially cause the re-emergence of gHAT. A stochastic model applied at a smaller spatial scale is more suitable for addressing issues related to chance events and their impact on EOT^11^.

A new oral drug to treat gHAT – fexinidazole – has now been approved for use in DRC, and is being utilised in the country. Despite the obvious advantages for patients, ease of transport and administration, it is not deemed suitable for use in individuals without parasitological case confirmation^24^, and hence is unlikely to greatly impact on transmission as part of a strategy. A second oral drug – acoziborole – hoped to be a safe single-dose cure, is under clinical trial and could, in principle, radically change the paradigm of diagnostic and treatment algorithms, especially in an AS setting^25^. The non-toxic compound used in acoziborole may allow mobile screening teams to “overtreat” rapid diagnostic test-positive, gHAT suspects without parasitological confirmation. Another important tool to measure elimination or detect re-emergence of gHAT as prevalence approaches zero is its diagnosis. In contrast to the need to detect as many cases as possible in epidemic and endemic situations, avoiding any false-positive results becomes more important when the prevalence is low. A promising new test with high specificity and sensitivity, iELISA, was newly developed for this purpose^26^. Different from the existing trypanolysis test, iELISA has lower requirements at both the laboratory and the technical skill levels and therefore is a better tool for post-elimination monitoring as endemic countries should be able to sustain it with little external financial and technical support. Mathematical modelling could be used to investigate the impact of potential diagnostic and treatment algorithms and predict the impact of such strategies using acoziborole and iELISA on EOT before they begin. These types of novel interventions could be particularly helpful as we approach the endgame for gHAT.

AS planning by PNLTHA-DRC is guided at a village level by WHO recommendations. These include stopping AS after three years of zero case detection and then switching to “reactive AS” when new cases arise in PS^27^. In the present study our four strategies were assumed to carry on indefinitely without any stopping, however the economic gains and health risks associated with cessation should be examined. A previous health economic analysis concluded that VC can be cost-effective at low willingness-to-pay thresholds per disability-adjusted life year averted in high-risk settings^28^. Taking account of the PNLTHA-DRC algorithm of reactive screening, a novel health economic analysis based on predicted model dynamics would allow for the examination of cost-effective strategies rather than the “preferred strategy” presented here based on a cruder ranking of “ambition”.

Looking across at other infections targeted for elimination, the enormity of the challenge ahead becomes apparent – with many of these programmes reaching ever lower levels of disease, but failing to meet elimination deadlines. Modelling in this study suggests that, even though elimination of gHAT in the near future may be epidemiologically feasible with current tools, its widespread, low-level persistence across the DRC could prove operationally challenging for achievement of the goal in the short-term. In many regions there is considerable uncertainty whether current interventions are sufficient to meet EOT in the next ten years, yet the prospect of intensifying strategies in dozens of health zones may pose a large, possibly insurmountable, burden on both financial and personnel resources. As further progress is made towards elimination of gHAT, it will become increasingly important to use data-driven methods to optimise the endgame pathway based on practical strategies and use these methods to quantify success.

## Methods

### Model

A previous modelling study showed that heterogeneous risk of human infections and participation in AS structures are essential to perform a good model fit to observed longitudinal human case data^8^. In this paper, we used a previously developed variant (“Model 4”) of the Warwick gHAT model^8,9,13^, which captures systematic non-participation of high-risk groups in the population – anecdotally believed to be working-age people spending time near tsetse habitat, and away from villages during active screening activities, to predict gHAT dynamics by considering transmission among humans, tsetse, and non-reservoir animals.

As illustrated in Figure 6, human hosts’ risk of infection categories are defined by their different contact rates with tsetse. High-risk humans (subscript *H*4) represent the working age males and are *r*-fold more likely to receive bites than low-risk humans (subscript *H*1). Any blood-meals taken upon “other” hosts do not result in infection. Both the proportion of low-risk humans (*k*_1_ from which we get the proportion of high-risk humans, *k*_4_ = 1 − *k*_1_) and the relative bites on high-risk humans (*r*) are fitted parameters in our model because we believe they would vary geographically. Tsetse select their blood-meal from one of the host types dependant upon innate feeding preference and relative host abundance. We assume tsetse preferentially feed on humans with a probability *f*_*H*_ which is taken to be 0.09^29^, if some other fixed value of *f*_*H*_ was used this would impact the fitting of the other model parameters, in particular *m*_*eff*_ (Supplementary Equation 1). In contrast to the assumption that low-risk humans randomly participate in active screening, high-risk humans are assumed to never participate. This participating structure is supported by data in previous model fits^13^. All infected individuals are assumed to exhibit treatment-seeking behaviour regardless of risk, however those in early stage infection have a much lower probability of seeking treatment compared to those with late stage infection. Thus passive detection is assumed to be dependent on their disease progression (slower rate to detection for stage 1, *η*_*H*_ compared to stage 2, *uγ*_*H*_), which includes the health zone-specific availability of fixed health facilities with gHAT diagnostics and underreporting. Tsetse bites are assumed to be taken on humans or non-reservoir animals. However, the non-reservoir animal species do not need to be explicitly modelled, i.e. this model variant does not include tsetse to non-human animal transmission. Complete mathematical descriptions are available in Supplementary Methods section with detailed updates in Supplementary Model Updates section.

**Figure 6.**
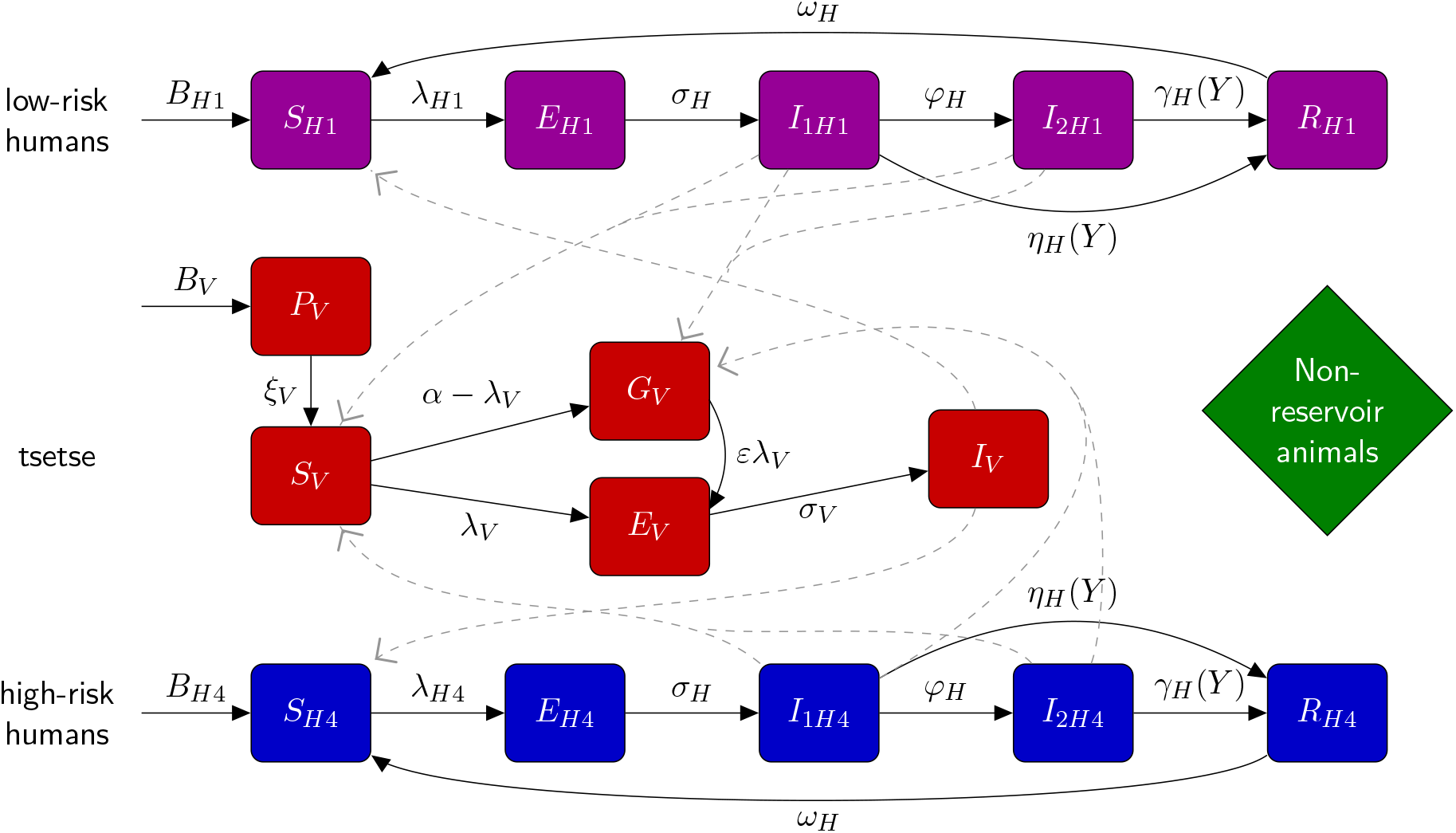
Illustration of compartmental gHAT model. Multi-host gHAT model is composed of one host species able to confer gHAT (humans), a further non-reservoir species (others) and tsetse. After incubation period, infected human hosts follow the progression which includes infectious stage 1 disease, *I*_1*H*_, infectious stage 2 disease, *I*_2*H*_, and non-infectious (due to hospitalisation) disease, *R*. Pupal stage tsetse, *P*_*V*_, emerge into unfed adults. Unfed tsetse are susceptible, *S*_*V*_, and following a blood-meal become either exposed, *E*_*V*_, or have reduce susceptibility to the trypanosomes, *G*_*V*_. Tsetse select their blood-meal from one of the host types dependant upon innate feeding preference and relative host abundance. High-risk humans are more likely to receive bites than low-risk humans. Any blood-meals taken upon “other” hosts do not result in infection. The transmission of infection between humans and tsetse is shown by grey paths. This figure is adapted from the original model schematic^8^.

### Fitting

In a previous publication we used the gHAT case data from the WHO HAT Atlas to fit the gHAT model to health zone specific trends from 2000–2016^13^. The data are aggregated by location, year and surveillance type, and location was defined by the available geolocation and geographical identifier information, while surveillance type was either active or passive screening^3,14,15^. Cases of gHAT are not reported across all of the DRC, and some health zones have very little case reporting or screening activities, and we do not include these locations in either fitting to the historical data or predictions of the future.For each health zone we fitted, we estimated model parameters that are likely to geographically vary across regions including: *R*_0_, *k*_1_, *r, η*_*H*_, *γ*_*H*_, *u* and *Spec* (Supplementary Table 2). The fitted model also takes into account previous advances in medical, diagnostic, and control systems; samples from posterior distributions of parameters were obtained by fitting to annual health zone-level data for the period 2000–2016 using an adaptive Metropolis-Hastings Markov chain Monte Carlo (MCMC) method^13^.

### Forward projections

Major changes during the data collection period include improvements to the PS systems in the former provinces of Bandundu and Bas Congo, improved active case confirmation via video recording of diagnostics in Mosango and Yasa Bonga in Bandundu from 2015, and implementation of large-scale VC in Yasa Bonga since mid-2015. Based on the continuation of the current PS system, we considered four strategies for projections from 2017 to 2050, which included different coverage of AS and whether or not to implement VC from 2020. As summarised in Table 1, AS is assumed to be either at the recent (2012–2016) mean level achieved or at the maximum level (maximum achieved during 2000–2016), and hence depends on the historical data in each health zone. For VC, a fixed effectiveness of 80% tsetse reduction after one year was used in the strategies with VC in all health zones except Yasa Bonga, where an effectiveness of 90% has been reported^16^. Other tsetse reductions (i.e. 60% and 90%) were considered in sensitivity analyses in Supplementary Fig. 1. Further model assumptions include: (1) in Yasa Bonga only strategies with VC are considered since VC was already in place before 2017; (2) video confirmation of parasitological diagnosis was included from 2018 in Bandundu health zones to avoid false positive diagnoses in AS; (3) automatic improvement of the diagnostic algorithms to 100% specificity outside Bandundu when the detected case numbers were close to the expected incidence of false positive detections.

The data set finished in 2016 and so forward projections were performed from 2017 to 2050, independently for each health zone. Parameter uncertainty was represented by 1,000 randomly selected sets of parameters from the health zone-specific posterior distributions from the model fitting. Observational uncertainty in predicted case numbers each year was considered by drawing ten random samples from the predicted mean dynamics for each set of parameters. In model outputs, 10,000 samples for observable variables such as active and passive cases, and related metrics were generated. On the other hand, unobservable variables like new infections and the year of EOT were predicted by the 1,000 model realisations (parameter uncertainty but no sampling uncertainty).

### Measuring elimination of transmission

As there is no direct way to observe EOT, WHO suggest a primary indicator of zero reported cases to measure the achievement of EOT^4,30^. However, the number of reported cases depends largely on the strength of medical interventions, so other methods to assess progress towards EOT are desirable to complement imperfect case indicators^15,31^.

Fortunately, mechanistic modelling provides the means to both infer and predict the unobservable transmission dynamics to assess EOT. Here, we calculated the number of underlying new infections each year in the epidemiological model. Unlike the discrete nature of populations, the outputs of deterministic models are continuous and whilst they can asymptote to zero they will never reach it. Therefore, to identify a realistic point at which EOT has been achieved, we introduced a proxy threshold (= 1) for annual new infections and assume that EOT is achieved when the number of new infections is below the threshold (Supplementary Model Updates section).

## Data Availability

The gHAT data were obtained from the WHO HAT Atlas and are subject to a data sharing agreement. Interested parties should apply to WHO in order to gain access to these data. Results for each health zone level projection can be viewed on the paper's companion website. Code and model outputs are available from the OSF.

https://hatmepp.warwick.ac.uk/projections/v1

https://osf.io/jza27/?view_only=d523cb1edf9c4828bc63cb197e5000b2

## Data availability

Data cannot be shared publicly because they were aggregated from the World Health Organisation’s HAT Atlas which is under the stewardship of the WHO. Data are available from the WHO (contact or visit https://www.who.int/trypanosomiasis_african/country/foci_AFRO/en/) for researchers who meet the criteria for access to confidential data. Data sharing is subject to WHO data-sharing policies and data-use agreements with the participating research centres.

## Code availability

The code used to simulate this work is available from: https://osf.io/jza27/?view_only=d523cb1edf9c4828bc63cb197e5000b2.

## Acknowledgments

The authors thank PNLTHA for original data collection, WHO for data access (in the framework of the WHO HAT Atlas^30^), and Cyrus Sinai and Nicole Hoff from UCLA Fielding School of Public Health for providing health zone level shapefiles (current versions can be found at https://data.humdata.org/dataset/drc-health-data). This work was supported, in whole or in part, by the Bill & Melinda Gates Foundation [OPP1177824, OPP1184344, OPP1156227, OPP1186851, OPP1155293]. Under the grant conditions of the Foundation, a Creative Commons Attribution 4.0 Generic License has already been assigned to the Author Accepted Manuscript version that might arise from this submission. This work was also supported by the Belgian Development Agency (ENABEL). The funders had no role in study design, data collection and analysis, decision to publish, or preparation of the manuscript.

## Author contributions

CH, REC, and KSR developed the software and performed the analyses. REC, CH, and PB visualised the results. REC, EMM, and CS analysed the data. KSR developed the methods. CH and KSR wrote the original draft. KSR, SEFS, and MJK conceptualised the study. All authors reviewed and approved the final version for publication.

## Additional Information

**Supplementary information** on mathematical modelling methodology is available.

