## Supplementary information for "Identifying regions for enhanced control of *gambiense* sleeping sickness in the Democratic Republic of Congo"

### Supplementary information: Identifying target regions for enhanced control of *gambiense* human African trypanosomiasis in the Democratic Republic of Congo

Ching-I Huang[a,b,*], Ronald E Crump[a,b,c,*], Paul Brown[a,b], Simon E F Spencer[a,d], Erick Mwamba Miaka[e], Chansy Shampa[e], Matt J Keeling[a,b,c], Kat S Rock[a,b]

[a]*Zeeman Institute for System Biology and Infectious Disease Epidemiology Research, The University of Warwick, Coventry, U.K.*

[b]*Mathematics Institute, The University of Warwick, Coventry, U.K.*

[c]*The School of Life Sciences, The University of Warwick, Coventry, U.K.*

[d]*The Department of Statistics, The University of Warwick, Coventry, U.K.*

[e]*Programme National de Lutte contre la Trypanosomiase Humaine Africaine (PNLTHA), Kinshasa, D.R.C.*

#### S1. Methods

This work is one of a series of studies on modelling and analyses on issues in the last mile of eliminating transmission of *gambiense* human African trypanosomiasis (gHAT) in the Democratic Republic of Congo (DRC). The series of studies starts with a model fitting paper[1] and to aid the reader of the present study, much of the same model information is provided here in the Methods section.

##### *S1.1. The compartmental gHAT model*

The gHAT model we considered in this study is a variant "Model 4" of the Warwick model presented in the literature[1,2,3,4]. gHAT infections among hosts are described by equation (S1). Human hosts are modelled by the SEIIRS model with two infectious compartments, stage 1 disease, $I_{1H}$, and stage 2 disease, $I_{2H}$. Vectors are modelled by using compartments for appropriately modelling tsetse when used in a host-vector model with disease[3]. Pupal stage tsetse, $P_V$, emerge into unfed susceptible adults, $S_V$, and following a blood-meal become either exposed, $E_V$, or have reduced susceptibility to the *Trypanosoma brucei gambiense* parasites, $G_V$ - this effect is known as the teneral phenomenon. Following an infection, tsetse have an extrinsic incubation period (EIP) before becoming onwardly infectious. To incorporate a more realistic EIP distribution, there are three exposed classes, $E_{1V}, E_{2V}, E_{3V}$, which result in a gamma-distributed EIP (rather than an exponential with only one).

In order to reduce the dimensionality of our ODE system (by one), the vector equations are non-dimensionalised using the scaling $N_H/N_V$, where $N_H$ is the total human population, and $N_V$ is the tsetse population size. This results in a new non-dimensionalised parameter, $m_{\text{eff}}$, which is $\dfrac{p_H N_V}{N_H}$ appearing in host equations ($p_H$ is the probability of a human being infected by a single infectious bloodmeal) and is referred to as the *effective vector density*.

The proportion of tsetse bites taken on low-risk and high-risk humans are $f_1$ and $f_4$, depending on the relative availability/attractiveness and the relative abundance of two risk groups. High-risk humans are assumed to be $r$-fold more

*These authors contributed equally to this work.

likely to receive bites, i.e. $s_1 = 1$ and $s_4 = r$. Therefore, $f_i$'s can be calculated using $f_i = \dfrac{s_i N_{Hi}}{\sum_j s_j N_{Hj}}$.

$$
\text{Humans}\left\{
\begin{aligned}
\frac{\mathrm{d}S_{Hi}}{\mathrm{d}t} &= \mu_H N_{Hi} + \omega_H R_{Hi} - \alpha m_{\text{eff}} f_i \frac{S_{Hi}}{N_{Hi}} I_V - \mu_H S_{Hi} \\
\frac{\mathrm{d}E_{Hi}}{\mathrm{d}t} &= \alpha m_{\text{eff}} f_i \frac{S_{Hi}}{N_{Hi}} I_V - (\sigma_H + \mu_H) E_{Hi} \\
\frac{\mathrm{d}I_{1Hi}}{\mathrm{d}t} &= \sigma_H E_{Hi} - (\varphi_H + \eta_H(Y) + \mu_H) I_{1Hi} \\
\frac{\mathrm{d}I_{2Hi}}{\mathrm{d}t} &= \varphi_H I_{1Hi} - (\gamma_H(Y) + \mu_H) I_{2Hi} \\
\frac{\mathrm{d}R_{Hi}}{\mathrm{d}t} &= \eta_H(Y) I_{1Hi} + \gamma_H(Y) I_{2Hi} - (\omega_H + \mu_H) R_{Hi}
\end{aligned}
\right.
$$

$$
\text{Tsetse}\left\{
\begin{aligned}
\frac{\mathrm{d}P_V}{\mathrm{d}t} &= B_V N_H - (\xi_V + \tfrac{P_V}{K}) P_V \\
\frac{\mathrm{d}S_V}{\mathrm{d}t} &= \xi_V \mathbb{P}(\text{survive pupal stage}) P_V - \alpha S_V - \mu_V S_V \\
\frac{\mathrm{d}E_{1V}}{\mathrm{d}t} &= \alpha\big(1 - f_T(t)\big) p_V \big(\textstyle\sum_i f_i \dfrac{(I_{1Hi} + I_{2Hi})}{N_{Hi}} + f_A \dfrac{I_A}{N_A}\big)(S_V + \varepsilon G_V) \\
&\qquad -(3\sigma_V + \mu_V + \alpha f_T(t)) E_{1V} \\
\frac{\mathrm{d}E_{2V}}{\mathrm{d}t} &= 3\sigma_V E_{1V} - (3\sigma_V + \mu_V + \alpha f_T(t)) E_{2V} \\
\frac{\mathrm{d}E_{3V}}{\mathrm{d}t} &= 3\sigma_V E_{2V} - (3\sigma_V + \mu_V + \alpha f_T(t)) E_{3V} \\
\frac{\mathrm{d}I_V}{\mathrm{d}t} &= 3\sigma_V E_{3V} - (\mu_V + \alpha f_T(t)) I_V \\
\frac{\mathrm{d}G_V}{\mathrm{d}t} &= \alpha(1 - f_T(t)) S_V \\
&\qquad -\alpha\Big(f_T(t) + (1 - f_T(t)) p_V \varepsilon \big(\textstyle\sum_i f_i \dfrac{(I_{1Hi} + I_{2Hi})}{N_{Hi}} + f_A \dfrac{I_A}{N_A}\big) G_V\Big) \\
&\qquad -\mu_V G_V
\end{aligned}
\right.
\tag{S1}
$$

*S1.2. Model assumptions and parameterisation*

Table S1 provides the estimates of fixed parameters available in the literature used in the previous gHAT model[1,2,3]. The parameters fitted during the model fitting are defined in Table S2. Posterior distributions for these parameters were based on the model fitting results[1].

*S1.3. Active screening*

Screening data is aggregated by year and the exact dates and frequencies of conducting AS are unknown, therefore some assumptions were made on when AS takes place. Our model assumed only low-risk humans participate in AS and used the ratio of assumed number of people screened ($N_{AS}$) and the number of low-risk humans ($k_1 N_H$) to decide the frequency of AS each year. When screening numbers were smaller than potential participants ($N_{AS} < k_1 N_H$), a single AS event was assumed to take place at the beginning of those years. On the other hand, multiple AS events were evenly distributed over the time of the corresponding years, i.e. a second AS event in July when $k_1 N_H < N_{AS} \leq 2k_1 N_H$; a second AS event in May and a third AS event in September when $2k_1 N_H < N_{AS} \leq 3k_1 N_H$; etc.

*S1.4. Formulation and parameterisation of improved passive detections in Bandundu and Bas Congo*

Previous analysis on provincial-level staged data (Lumbala *et al.* for 2000–2012[16] and WHO HAT Atlas data 2015–2016[17]) indicated that improved passive detection has happened across former Bandundu province and in former Bas

**Table S1: Model parameterisation (fixed parameters).** Notation, a brief description, and the used values for fixed parameters.

| Notation | Description | Value | |
|---|---|---|---|
| $N_H$ | Total human population size in 2015 | Fixed for each health zone | [5] |
| $\mu_H$ | Natural human mortality rate | $5.4795\times10^{-5}$ days$^{-1}$ | [6] |
| $B_H$ | Total human birth rate | $= \mu_H N_H$ | |
| $\sigma_H$ | Human incubation rate | 0.0833 days$^{-1}$ | [7] |
| $\varphi_H$ | Stage 1 to 2 progression rate | 0.0019 days$^{-1}$ | [8,9] |
| $\omega_H$ | Recovery rate or waning-immunity rate | 0.006 days$^{-1}$ | [10] |
| Sens | Active screening diagnostic sensitivity | 0.91 | [11] |
| $B_V$ | Tsetse birth rate | 0.0505 days$^{-1}$ | [3] |
| $\xi_V$ | Pupal death rate | 0.037 days $^{-1}$ | |
| $K$ | Pupal carrying capacity | $= 111.09N_H$ | [3] |
| $\mathbb{P}$(pupating) | Probability of pupating | 0.75 | |
| $\mu_V$ | Tsetse mortality rate | 0.03 days$^{-1}$ | [7] |
| $\sigma_V$ | Tsetse incubation rate | 0.034 days$^{-1}$ | [12,13] |
| $\alpha$ | Tsetse bite rate | 0.333 days$^{-1}$ | [14] |
| $p_V$ | Probability of tsetse infection per single infective bite | 0.065 | [7] |
| $\varepsilon$ | Reduced non-teneral susceptibility factor | 0.05 | [2] |
| $f_H$ | Proportion of blood-meals on humans | 0.09 | [15] |
| $\text{disp}_{\text{act}}$ | Overdispersion parameter for active detection | $4\times10^{-4}$ | [1] |
| $\text{disp}_{\text{pass}}$ | Overdispersion parameter for passive detection | $2.8\times10^{-5}$ | [1] |

[1] Value of $B_V$ is chosen to maintain constant population size without interventions.
[2] Value of $K$ is chosen to reflect the observed bounce back rate.

**Table S2: Model parameterisation (posteriors of fitted parameters).** Notation, a brief description, and representative percentiles of the posterior distributions for fitted parameters.

| Notation | Description | Posterior (median [95% CI]) Kwamouth | Tandala |
|---|---|---|---|
| $R_0$ | Basic reproduction number (NGM approach) | 1.09 [1.06, 1.14] | 1.009 [1.006, 1.014] |
| $r$ | Relative bites taken on high-risk humans | 6.61 [3.15, 10.75] | 2.04 [1.30, 4.26] |
| $k_1$ | Proportion of low-risk people | 0.90 [0.82, 0.95] | 0.95 [0.85, 0.99] |
| $\gamma_H^{\text{pre}}$ | Pre-1998 treatment rate from stage 2 (days$^{-1}$) | $1.72 \times 10^{-3}$ $[0.38, 4.88] \times 10^{-3}$ | $2.53 \times 10^{-3}$ $[1.03, 7.15] \times 10^{-3}$ |
| $\eta_H^{\text{post}}$ | Post-1998 treatment rate from stage 1 (days$^{-1}$) | $1.24 \times 10^{-4}$ $[0.60, 2.74] \times 10^{-4}$ | $2.74 \times 10^{-4}$ $[1.11, 4.99] \times 10^{-4}$ |
| $\gamma_H^{\text{post}}$ | Post-1998 treatment rate from stage 2 (days$^{-1}$) | $1.88 \times 10^{-3}$ $[0.46, 5.42] \times 10^{-3}$ | $3.60 \times 10^{-3}$ $[1.72, 8.98] \times 10^{-3}$ |
| Spec | Active screening diagnostic specificity | 0.9991 [0.9987, 0.9997] | 0.9998 [0.9997, 0.9999] |
| $u$ | Proportion of stage 2 passive cases reported | 0.27 [0.18, 0.40] | 0.39 [0.29, 0.51] |
| $d_{\text{change}}$ | Midpoint year for passive improvement | 2005.8 [2004.4, 2007.3] | – |
| $\eta_{H_{\text{amp}}}$ | Relative improvement in passive stage 1 detection rate | 2.52 [0.92, 5.46] | – |
| $\gamma_{H_{\text{amp}}}$ | Relative improvement in passive stage 2 detection rate | 0.51 [0.24, 0.97] | – |
| $d_{\text{steep}}$ | Speed of improvement in passive detection rate (years$^{-1}$) | 0.94 [0.68, 1.29] | – |

[1] See equation (S2) for improved passive detections formulated by $d_{\text{change}}$, $\eta_{H_{\text{amp}}}$, $\gamma_{H_{\text{amp}}}$ and $d_{\text{steep}}$.

Congo province[1]. Logistic functions shown in equation (S2) were used to formulate the improved passive detections in Bandundu and Bas Congo in year $Y$.

$$\begin{aligned} \eta_H(Y) &= \eta_H^{\text{post}}\left[1+\frac{\eta_{H_{\text{amp}}}}{1+\exp\left(-d_{\text{steep}}(Y-d_{\text{change}})\right)}\right], \\ \gamma_H(Y) &= \gamma_H^{\text{post}}\left[1+\frac{\gamma_{H_{\text{amp}}}}{1+\exp\left(-d_{\text{steep}}(Y-d_{\text{change}})\right)}\right]. \end{aligned} \tag{S2}$$

Parameter definitions and their posteriors are provided in Table S2. N.B. It was assumed that improvements in both stages shared the same midpoint year and speed of improvement within a health zone. However, the amplitude of variation in each health zone came from the fitting of health-zone-specific data.

*S1.5. Formulation and parameterisation of additional tsetse mortality under vector control measures*

The function which describes the probability of both hitting a target and dying is time dependent (days) from when the targets where placed:

$$f_T(t) = f_{\text{max}}\left(1-\frac{1}{1+\exp(-0.068(\text{mod}(t,182.5)-127.75))}\right), \tag{S3}$$

and $f_{\text{max}}$ is chosen such that the tsetse population after one year is at the observed/assumed percentage reduction. For the simplified model this is given by $f_{\text{max}} = 0.0305$ for a 60% reduction, $f_{\text{max}} = 0.0525$ for an 80% reduction, and $f_{\text{max}} = 0.0750$ for a 90% reduction.

*S1.6. Simulations performed*

Simulations were performed based on 1,000 model realisations. Observation uncertainty was considered by drawing ten random samples from the predicted mean dynamics for each set of parameters. A beta-binomial distribution in which an overdispersion parameter $\rho$ was introduced to the binomial distribution was used to account for larger variance than the binomial. The probability of obtaining $m$ successes out of $n$ trials with probability $p$ and overdispersion parameter $\rho$ is

$$\text{BetaBin}(m; n, p, \rho) = \frac{\Gamma(n+1)\Gamma(m+a)\Gamma(n-m+b)\Gamma(a+b)}{\Gamma(n-m+1)\Gamma(n+a+b)\Gamma(a)\Gamma(b)}, \tag{S4}$$

where $a = p(1/\rho - 1)$ and $b = a(1-p)/p$.

Main observable outputs including active and passive cases each year were predicted by 10,000 samples. Unobservable outputs, such as new infections and the year of EOT, were predicted directly from the 1,000 model realisations without sampling (parameter uncertainty but no observation uncertainty). Our model also has the capability of outputting unreported deaths and person years spent in stage 1 and stage 2.

*S1.7. Uncertainty*

Model predictions propagate parameter and observation uncertainty (see above). We tried to represent this uncertainty in a variety of ways:

- Time series box plots were used to display statistical summaries of model predictions – the median (the middle line in each box), the lower and upper quartiles (the edges of each box showing 50% prediction intervals) and 95% prediction intervals (extended whiskers containing the middle 95% of outputs).
- The median of YEOT was used to indicate the estimated elimination year for a series of model predictions because neither extreme values (outliers) nor truncation of simulation will affect the estimates.
- Probability of EOT by 2030 (or any given year between 2020 and 2040 in GUI) was calculated as the proportion of model realisations that achieve EOT by 2030 or the given year. Its continuous spectrum shows how likely we are to achieve EOT by a particular year and highlights regions we are most uncertain about with respect to meeting this goal.
- Sensitivity analysis for VC was performed to provide additional assessment of the VC reduction assumption (see results below).

##### *S1.8. Proxy for EOT*

The gHAT model we used here is a deterministic model described by ODEs with transition rates between compartments. In the deterministic model, variables such as new infections, new cases and deaths can be non-integer and their values are continuous. The stochastic model, on the other hand, has dynamics driven by randomly occurring events with associated probabilities and its variables capture the discrete nature of the population. Despite good agreements on the mean dynamics in both models, even at very low prevalence, the dynamics at the endgame are different. Because of the continuous nature of deterministic dynamics, the number of infected people asymptotes to zero rather than reaching it unlike the stochastic model. In this paper, an artificial EOT threshold i.e. one new infection per health zone per year was applied to new infections to determine whether EOT has been achieved or not. Other values of EOT thresholds, such as one new infection per 100,000 or per 1,000,000 people per year, can be found in the literature[18,19]. Large variation in EOT threshold highlights the difficulty in choosing a proper threshold reflecting the reality. More detailed comparison between stochastic and deterministic model variants will be needed in the future to ensure robustness of year of EOT estimates arising from such a proxy threshold.

#### S2. Results

##### *S2.1. Sensitivity analysis of the effectiveness of vector control*

The reported annual reductions in tsetse populations of vector control (VC) range from 80% to 99%[4,20,21,22]. The reductions are highly variable between locations due to differences in accessibility and the effectiveness of target deployments. In addition to the default tsetse reduction assumed here (80%), a conservative and a high but achievable reduction, 60% and 90%, are considered in the sensitivity analysis of VC. As shown in figure S1, the sensitivity analysis shows that the effectiveness of VC has a direct impact on the numbers of underlying new infections and leading to a delay to EOT when VC effectiveness is low.

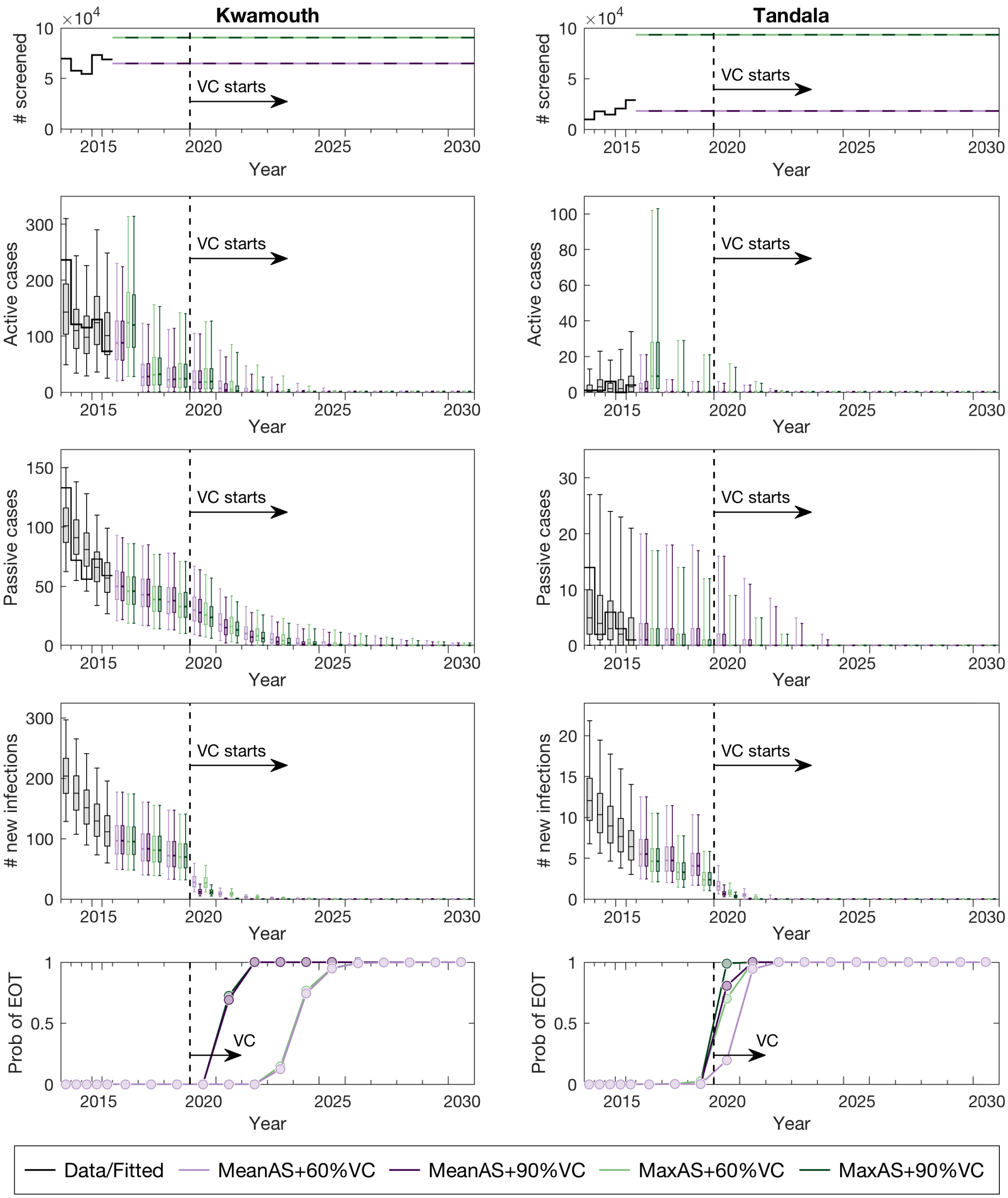


**Figure S1: Sensitivity analysis of vector control.** The reductions in tsetse populations depend on the accessibility and the effectiveness of target deployments. A conservative reduction (60%) and a high but achievable reduction (90%) are considered in the sensitivity analysis of VC. VC efficiently stops the transmission from tsetse to humans and therefore greatly reduces the number of new infections. The decreases in new infections reflect the effectiveness of vector control, i.e. 90% VC (dark purple and dark green) has fewer expected new infections than 60% VC (light purple and light green). As a result, time lags in achieving EOT for the less effective VC strategy are shown in both high-risk and low-risk health zones.

#### S3. Model Updates

Variations on this "Warwick gHAT model" have been published previously (see Section 1.1). Key differences between this model and previous versions (notably[19]) have been described in the study when the model was fitted to data[1] and are listed again here:

- Improved passive detection: Improvement due to the introduction of CATT test ($\eta_H^{\mathrm{pre}} = 0 \mapsto \eta_H^{\mathrm{post}}$ and $\gamma_H^{\mathrm{pre}} = b_{\gamma_H^{\mathrm{pre}}} \gamma_H^{\mathrm{post}} \mapsto \gamma_H^{\mathrm{post}}$ in Crump *et al*[1]) is considered in 1998 in the entire DRC as well as gradual improvements of stage 1 and stage 2 passive detection rates over time (see Subsection 1.4 above) are taken into account in Bandundu and Bas Congo provinces.

- MSF AS algorithm: The results of CATT 1:32, which has a higher sensitivity (MSF sensitivity = 0.95 in contrast to PNLTHA sensitivity = 0.91) and a lower fitted specificity ($= b_{\mathrm{specificity}} \times \mathrm{specificity}$ with targeted mean = 0.991 in Crump *et al*[1]) than the PNLTHA-DRC algorithm, were used for case reporting and treatments in Orientale province in active screenings performed by MSF before 2013.

- Perfect specificity in AS: Video confirmation was introduced in Mosango and Yasa Bonga since 2015 and therefore the specificity in active screenings became 1 since then. Perfect specificity is also assumed in the rest of Bandundu from 2018.

- Overdispersion in case detections: Observation uncertainty is considered by adding overdispersion into case detections (see Subsection 1.6 above). To avoid overfitting, the overdispersion parameters were manually tuned to be appropriate for a health zone level fit, and thereafter left fixed across MCMC runs in Crump *et al*[1].

- EOT threshold: A proxy threshold (= 1) of new infection per health zone per year is used to identify when EOT has been reached within this deterministic framework. Previous versions of this model have used a variety of proxy thresholds, typically fewer than one new infection per 100,000 per year[18,19].

- No transmission after achieving EOT: Unlike previous Warwick gHAT model variants, once the EOT threshold is met we set transmission to zero in subsequent years and therefore no further new infections is possible. Previously infected people can still be identified and reported (although there are typically extremely few reported cases at that point).

#### S4. PRIME-NTD criteria

It has been recommended that good modelling practises should meet the five key principles relating to communication, quality and relevance of analyses – known as Policy-Relevant Items for Reporting Models in Epidemiology of Neglected Tropical Diseases (PRIME-NTD)[23]. We present how these PRIME-NTD criteria have each been addressed in Table S3.

**Table S3:** PRIME-NTD criteria fulfillment. We summarise how the NTD Modelling Consortium's "5 key principles of good modelling practice" have been met in the present study.

| Principle and what has been done to satisfy the principle? | Where in the manuscript is this described? |
|---|---|
| **1. Stakeholder engagement**<br>This study was lead by modellers and guided by members of the national sleeping sickness control programme in DRC (PNLTHA-DRC) – coauthors E Mwamba Miaka and S Chancy. PNLTHA-DRC have contributed to improved modeller understanding of the epidemiological data and changes to the programme over time and in different geographic regions, both of which impacted model fitting over several rounds of revision (via in-person meetings and email). The GUI (and several variants of it) was designed in conjunction with PNLTHA-DRC to improve communication of the modelling outputs to non-modellers. It has been refined through various in-person meetings with different collaborators with the goal of providing understandable, policy-relevant outputs as well as scientific communication; over 20 non-modellers have had opportunities to interact with and provide feedback on the GUI during development. | Authorship list |
| **2. Complete model documentation**<br>Full model (including the fitting code) and documentation are available through Open Science Framework (OSF). The model is fully described in this SI and in the fitting study of Crump *et al*[1]. | See Section S1 above in this document and access the code via `ProjOSF` |
| **3. Complete description of data used**<br>The data used for fitting were described in detail in Crump *et al*[1]. Posteriors used for the projections presented here are available on our OSF page. | Posteriors available at `FittingOSF` |
| **4. Communicating uncertainty**<br>*Structural uncertainty*:<br>The variant of the model presented here ("Model 4") was chosen as it had good support compared to other plausible model structures when fitting to data sets from Yasa Bonga and Mosango health zones in DRC[2] and in the Mandoul focus, Chad[4]. | See Methods section in main text. |
| *Parameter uncertainty*:<br>In the fitting study[1] key model parameters were fitted in an MCMC framework by utilising regional data, we used these posterior parameter sets to simulate forward projections in the present study. Sensitivity analysis was performed on the assumed impact of tsetse reduction through vector control (VC) with 60% and 90% reductions presented in addition to the default 80% assumption. | See Methods section in main text and Crump *et al*[1], and posterior files are available at `FittingOSF`. Sensitivity analysis for VC in Section S2.1 above and in `GUI`. |
| *Prediction uncertainty*:<br>Here, we represent uncertainty in our results in various ways - (i) by providing box and whisker plots for predictions (median, 50% and 95% prediction intervals), (ii) by utilising probability maps (likelihood of meeting EOT goal by 2030) in addition to median year of EOT maps, and (iii) by providing prediction intervals for EOT years in the hoover feature of our GUI. | See Figures 1 and 3 in the main text and maps in `GUI`. |
| **5. Testable model outcomes**<br>Predictions presented here include measurable, and routinely reported outcomes such as active and passive case reporting by year. Outputs in the GUI can be compared to new case data as it becomes avaliable. Our predictions are dependant on the coverage of active screening each year and are expected to perform better when the actual screening coverage is similar to that assumed for predicting. In the future we plan to validate the predictions based on at least two years of new case data to assess model performance. | Predictions presented in main text Figure 1 and in the `GUI`. Model code and posteriors which could be used for validation are available at `ProjOSF` and `FittingOSF`. |

[1] Hyperlink `ProjOSF` with full address: `https://osf.io/jza27/?view_only=526344c12324492083db1e49c76136af`.
[2] Hyperlink `FittingOSF` with full address: `https://osf.io/ck3tr/?view_only=526344c12324492083db1e49c76136af`.
[3] Hyperlink `GUI` with with full address: `https://hatmepp.warwick.ac.uk/projections/v1`

14 World Health Organization & WHO Expert Committee on the Control and Surveillance of Human African Trypanosomiasis (2013: Geneva, Switzerland). Control and surveillance of human African trypanosomiasis: report of a WHO expert committee. WHO technical report series; no. 984. World Health Organization; 2013.

15 Clausen PH, Adeyemi I, Bauer B, Breloeer M, Salchow F, Staak C. Host preferences of tsetse (Diptera: Glossinidae) based on bloodmeal identifications. Medical and Veterinary Entomology. 1998;12:169–180.

16 Lumbala C, Simarro PP, Cecchi G, Paone M, Franco JR, Kande Betu Ku Mesu V, et al. Human African trypanosomiasis in the Democratic Republic of the Congo: disease distribution and risk. International Journal of Health Geographics. 2015;14:20.

17 Simarro PP, Cecchi G, Jannin JG. The Atlas of human African trypanosomiasis: a contribution to global mapping of neglected tropical diseases. International Journal of Health Geographics. 2010;9:57.

18 Rock KS, Ndeffo-Mbah ML, Castaño S, Palmer C, Pandey A, Atkins KE, et al. Assessing strategies against gambiense sleeping sickness through mathematical modeling. Clinical Infectious Diseases. 2018;66:S286–S292.

19 Castaño MS, Ndeffo-Mbah ML, Rock KS, Palmer C, Knock E, Mwamba Miaka E, et al. Assessing the impact of aggregating disease stage data in model predictions of human African trypanosomiasis transmission and control activities in Bandundu province (DRC). PLOS Neglected Tropical Diseases. 2020;14:e0007976.

20 Courtin F, Camara M, Rayaisse JB, Kagbadouno M, Dama E, Camara O, et al. Reducing human-tsetse contact significantly enhances the efficacy of sleeping sickness active screening campaigns: a promising result in the context of elimination. PLOS Neglected Tropical Diseases. 2015;9:e0003727.

21 Tirados I, Esterhuizen J, Kovacic V, Mangwiro TNC, Vale GA, Hastings I, et al. Tsetse control and gambian sleeping sickness; implications for control strategy. PLOS Neglected Tropical Diseases. 2015;9:1–22.

22 Tirados I, Hope A, Selby R, Mpembele F, Miaka EM, Boelaert M, et al. Impact of tiny targets on *Glossina fuscipes quanzensis*, the primary vector of human African trypanosomiasis in the Democratic Republic of the Congo. PLOS Neglected Tropical Diseases. 2020;14:1–20.

23 Behrend MR, Basáñez MG, Hamley JID, Porco TC, Stolk WA, Walker M, et al. Modelling for policy: The five principles of the Neglected Tropical Diseases Modelling Consortium. PLOS Neglected Tropical Diseases. 2020;14:e0008033.
